# The Local Initiative For Emergency Blood (LIFE-Blood) Study: A mixed-methods, single center exploration of civilian walking blood bank need, feasibility, and safety in a low-resource blood desert

**DOI:** 10.1101/2025.05.23.25327506

**Authors:** Nakul Raykar, Tecla Chelagat, Linda S. Barnes, Nikathan Kumar, Vanitha Raguveer, Cindy Makanga, Abdirahman Musa, Wendy Williams, Gilchrist Lokoel, Stephen Omondi Onyango, Edward Mutebi, Epem Esekon, Andrew P. Cap, Meghan Delaney, Marisa Del Signore, Tanujit Dey, Saksham Gupta, Pratap Kumar, Shreenik Kundu, Fleming Mathew, Juan Carlos Puyana, Nobhojit Roy, Riya Sawhney, Elizabeth Stoeger, Neil Thivalapill, Avery A. Thompson, Alejandro Munoz Valencia, Caroline Wesonga Wangamati

## Abstract

**Background:** Billions live in blood deserts, regions where there is effectively no access to blood transfusions. Lodwar County Referral Hospital, set in a rural, low-resource area in northwestern Kenya, is one such blood desert. A walking blood bank is a point-of-care system of emergency transfusion when banked blood is unavailable. The objective of this study was to evaluate the need, feasibility, and safety of a walking blood bank to address blood unavailability in a low-resource, civilian setting.

**Study Design and Methods:** This was a mixed method study. Blood need was determined through chart review from April to July 2022. Feasibility was evaluated through qualitative interviews with hospital stakeholders. Safety was established by comparing diagnostic performance of rapid diagnostic testing to standard-of-care assays for transfusion transmitted infections (HIV, HBV, HCV, syphilis).

**Results:** Stockouts occurred 40 of 126 days (32%). Stakeholders acknowledged blood was frequently unavailable and it was sometimes necessary to perform emergency transfusion using rapid tests to screen blood in order to save a life, despite uncertainty over their performance to detect transfusion infections. Overall transfusion infection prevalence in donors was 5.4%. Rapid testing had a 99.2% (p=0.01) negative predictive value when compared against standard-of-care laboratory-based tests.

**Discussion:** A walking blood bank using rapid testing may serve as a stopgap measure to address the extensive burden of hemorrhagic shock and severe anemia in the world’s lowest resource, civilian settings, where patients and their providers struggle without timely access to blood through a blood bank.

## Introduction

Billions live in “blood deserts,” regions of the world without any access to blood, where healthcare facilities are hours to days away from the nearest, stocked blood bank.^1,2^ Most blood deserts exist in low- and middle-income countries (LMICs), resulting in uncounted death, disability, and delays in care due to hemorrhage and severe anemias.^1,2^

When faced with bleeding patients and no access to transfusion, practitioners in blood deserts have historically had limited options: let patients die because of lack of blood or ‘transfer’ patients to another facility, knowing they are unlikely to make it. Another lesser used option would be to mobilize a walking blood bank (WBB)^3^. While a WBB can take on many forms depending on context, in general, it involves 1.) emergency mobilization of available local donors (often family, healthcare workers, or community members); 2.) the use of point-of-care, rapid diagnostic testing (RDT) instead of more extensive laboratory-based methods for blood screening for transfusion transmissible infection (TTI); and 3.) transfusion of minimally processed fresh whole blood after appropriate compatibility testing.^4,5^ Because of fears that RDT was less effective in detecting TTIs compared to laboratory-based enzyme immunoassay (EIA), particularly in the context of the HIV/AIDS epidemic and high rates of hepatitis in many LMICs, RDT for blood transfusion screening was strongly discouraged or explicitly outlawed in many countries, effectively ruling out the possibility of establishing WBBs.^6–9^

The past two decades, however, have seen major advances in WBB emergency transfusion strategies, and in RDT.^10^ First, militaries have adopted and refined the WBB strategy for transfusion needs in austere, battlefield environments.^11,12^ The US military, for example, has transfused thousands of units of fresh whole blood with excellent safety profiles.^13–16^ Second, WBBs have been used in high-income civilian environments to address mass casualty events and or as a preparedness measure to meet emergency transfusion needs.^17–22^ Importantly, RDT quality has improved dramatically, with the latest generation of testing kits rivaling performance of more expensive EIA, which is the TTI screening standard in many LMICs.^23^

However, despite these developments, WBB use remains scarce, possibly due to significant knowledge gaps in the need for, feasibility, and safety of implementing a WBB in a civilian, LMIC context. For example, what is the actual availability and insufficiency of blood at single facilities in blood deserts? Do local providers and hospital staff believe these shortages would be amenable to improvement with a walking blood bank? What are other barriers and facilitators of WBB implementation that exist in these environments? Furthermore, in spite of excellent manufacturer-reported performance in TTI detection, perspectives are common amongst blood transfusion authorities (and providers) that RDT poses major TTI transmission risks to patients due to inferior sensitivity for TTI, especially under harsh environmental conditions characteristic of many LMIC blood deserts. What is the actual performance of RDT compared to standard-of-care EIA in a hot environment with long transport times for consumable supply chains? We aimed to close these knowledge gaps and evaluate the need, feasibility, and safety of a WBB to address severe, chronic blood insufficiency in one of the world’s most difficult civilian healthcare delivery environments: the blood (and physical) desert of Turkana County, Kenya.

## Study Design and Methods

### Study Setting

Turkana County is Kenya’s largest county by landmass and home to 1.2 million people, many of whom live nomadic lifestyles in remote settings.^24^ The region has one referral hospital based in the capital city of Lodwar known as Lodwar County Referral Hospital (LCRH). LCRH has a blood bank that collects, stores, and distributes blood but does not have EIA-based TTI screening capabilities on-site. Therefore, LCRH sends samples of collected donor blood to a Regional Blood Transfusion Service (RBTS) center 360 km away for TTI screening. Blood collected from a donor will not be available for transfusion for 3-4 days, on average, until the RBTS has screened blood and communicated back to LCRH. RDT for HIV, hepatitis, and syphilis are readily available at LCRH for general infection screening but are not authorized for use in transfusion contexts given concerns over sensitivity for TTIs and TTI transmission through use of blood screened with RDTs instead of EIA.

### Study Design

We designed a mixed methods study to assess the need, feasibility, and safety of a WBB (Table 1) in Turkana, Kenya. This study was performed in collaboration with a broader study assessing blood system optimization in Kenya.^25^ Need for blood transfusion was determined through quantitative evaluation of blood demand (transfusion requests) and usage (request fulfillment) at LCRH and qualitative interviews. Feasibility and the potential for standardization was evaluated through qualitative interviews. Safety was evaluated by assessing RDT diagnostic performance for TTI detection, compared to the standard-of-care.

**Table 1.**
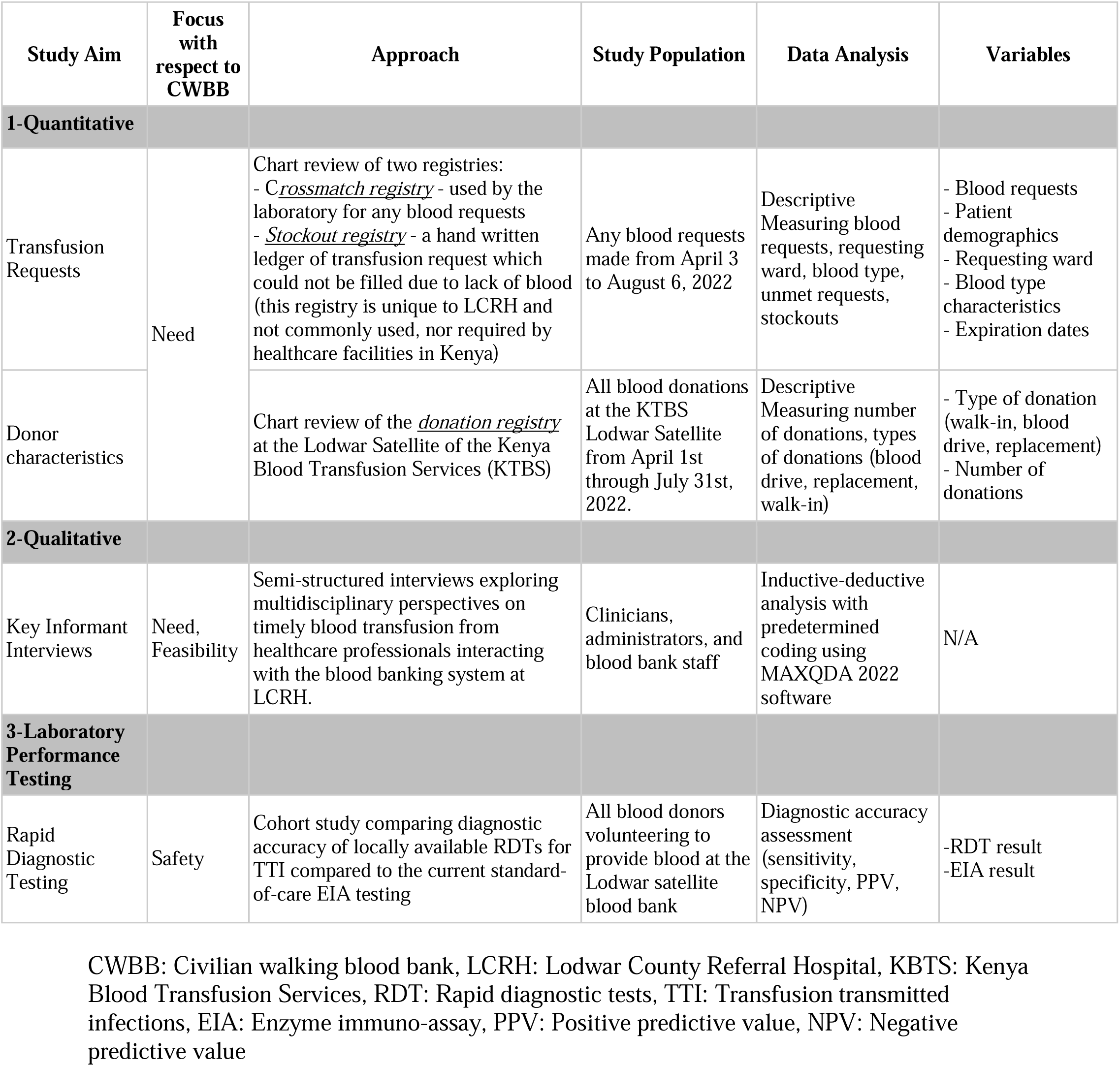
Methods Summary.

| Study Aim | Focus with respect to CWBB | Approach | Study Population | Data Analysis | Variables |
| --- | --- | --- | --- | --- | --- |
| 1-Quantitative |  |  |  |  |  |
| Transfusion Requests | Need | Chart review of two registries:<br>- *Crossmatch registry* - used by the laboratory for any blood requests<br>- *Stockout registry* - a hand written ledger of transfusion request which could not be filled due to lack of blood (this registry is unique to LCRH and not commonly used, nor required by healthcare facilities in Kenya) | Any blood requests made from April 3 to August 6, 2022 | Descriptive<br>Measuring blood requests, requesting ward, blood type, unmet requests, stockouts | - Blood requests<br>- Patient demographics<br>- Requesting ward<br>- Blood type characteristics<br>- Expiration dates |
| Donor characteristics |  | Chart review of the *donation registry* at the Lodwar Satellite of the Kenya Blood Transfusion Services (KTBS) | All blood donations at the KTBS Lodwar Satellite from April 1st through July 31st, 2022. | Descriptive<br>Measuring number of donations, types of donations (blood drive, replacement, walk-in) | - Type of donation (walk-in, blood drive, replacement)<br>- Number of donations |
| 2-Qualitative |  |  |  |  |  |
| Key Informant Interviews | Need, Feasibility | Semi-structured interviews exploring multidisciplinary perspectives on timely blood transfusion from healthcare professionals interacting with the blood banking system at LCRH. | Clinicians, administrators, and blood bank staff | Inductive-deductive analysis with predetermined coding using MAXQDA 2022 software | N/A |
| 3-Laboratory Performance Testing |  |  |  |  |  |
| Rapid Diagnostic Testing | Safety | Cohort study comparing diagnostic accuracy of locally available RDTs for TTI compared to the current standard-of-care EIA testing | All blood donors volunteering to provide blood at the Lodwar satellite blood bank | Diagnostic accuracy assessment (sensitivity, specificity, PPV, NPV) | -RDT result<br>-EIA result |

CWBB: Civilian walking blood bank, LCRH: Lodwar County Referral Hospital, KBTS: Kenya Blood Transfusion Services, RDT: Rapid diagnostic tests, TTI: Transfusion transmitted infections, EIA: Enzyme immuno-assay, PPV: Positive predictive value, NPV: Negative predictive value

### Quantitative Assessment of Blood Donation, Usage, Collection

We reviewed LCRH blood bank and laboratory registries to characterize blood availability and sufficiency (Table 1). A “crossmatch” register, maintained by the LCRH laboratory, recorded results of a crossmatch every time it was performed, just prior to dispatch of the unit to a patient ward. A separate “stockout” register used by LCRH laboratory staff noted blood requests that went unfilled due to stockout. We collected transfusion request and dispatch data in the two registries for four months. We additionally examined the donation registry over the same period, detailing the quantity and sources of blood donations at LCRH’s blood bank. Replacement donations refer to donations by patients’ family or friends to restock blood that will be used for the patient. These hand-written registries were anonymized, uploaded electronically, and collated to create a combined dataset. We ran descriptive statistics quantifying blood needs using Excel (Microsoft).

### Qualitative Interviews

We conducted individual key informant interviews with LCRH staff and leadership in English. Ten participants were recruited and consented using purposive sampling, based on roles in blood ordering and administration. Interview guides encompassed perspectives on local blood availability and practices when no banked blood was present. A second interview guide was created specifically for blood banking staff, containing additional questions related to testing and screening of blood. Interviews were recorded, de-identified, and transcribed with Otter.AI (2022). We analyzed transcripts using MAXQDA 2022 (VERBI Software, 2021). All available participants (n=7) and an additional new stakeholder engaged in a focus group discussion to provide feedback on emerging themes. Two groups of four participants reflected on two real-world patient scenarios while a researcher simultaneously recorded considerations (Supplement 3). Following this process, researchers revised the codebook a final time and analyzed transcripts identifying new codes and refining final themes.

### Safety

We assessed the diagnostic performance of RDT compared to the Kenyan standard-of-care, EIA, for TTIs. Testing included HIV, Hepatitis B (HBV) and C (HCV), and syphilis. RDT used in this study, along with manufacturer-reported sensitivities, were as follows: HIV: Abbott 7D2342, 99.91% sensitivity; HBV: Wondfo W3-S, 96.2% sensitivity; HCV: OnSite r0023S, 98.7% sensitivity; Syphilis: Wondfo W34-S4P, 100% sensitivity. All RDT was available through standard procurement practices at LCRH. All collected blood donations had samples sent for EIA testing at the RBTS, per standard process. In addition, we obtained additional samples from each donor blood unit. An LCRH laboratory technician performed RDT; results were logged in an electronic record. EIA and RDT results were compared. When EIA and RDT results were discordant (i.e., negative RDT and positive EIA), the RDT was repeated on a retained sample of blood.

To study the limit of detection of RDT, serial dilution testing with confirmed TTI-positive samples was completed. We created reference samples, using a private external laboratory to quantify viral loads and/or optical densities of the EIA test in positive samples for each TTI except for syphilis. Since syphilis is a bacterium, only optical density of the immunoassay was measured. Each reference sample was split into six aliquots before undergoing twelve levels of 1:1 dilution with normal saline. We ran RDT for each aliquot at every level of dilution and recorded the results.

### Ethics Approval and Role of Funding Source

This study received ethics approval from Strathmore University, Nairobi, Kenya (#SU-IERC1296/22); Mass General Brigham, Boston, MA (#2022P001253, 2022P001254); and the National Commission on Science, Technology, and Innovation, Nairobi, Kenya (#982141). The study was performed with funding from the Gillian Reny Stepping Strong Center for Trauma Innovation at the Brigham and Women’s Hospital in Boston, MA. The funder had no role in study design, data collection, data analysis, data interpretation, or writing of the manuscript.

## Results

### Quantitative Assessment of Blood Donation, Demand, Usage

Over the study period (126 days), 486 requests for blood transfusion were submitted to the LCRH blood bank. Of ten hospital wards, surgery (146, 30.04%), medicine (134, 27.57%), and maternity (87, 17.90%) submitted the most requests (Supplemental Table 1). Stockouts – requests not filled because compatible, screened blood was unavailable – occurred on 40 (31.74%) non-consecutive days. During each stockout day, the number of unmet requests ranged from 1 to 5 units (Figure 1). The blood bank filled 409 (84.15%) requests, all with whole blood. There was no local capacity for componentization of blood products during the study period. The average requests per month was 184 (range=160-217). The longest continuous stockout periods lasted four days; occurring twice during the study period. Two-day stockouts occurred nine times. The remaining stockouts were single days (14 occurrences).

**Figure 1.**
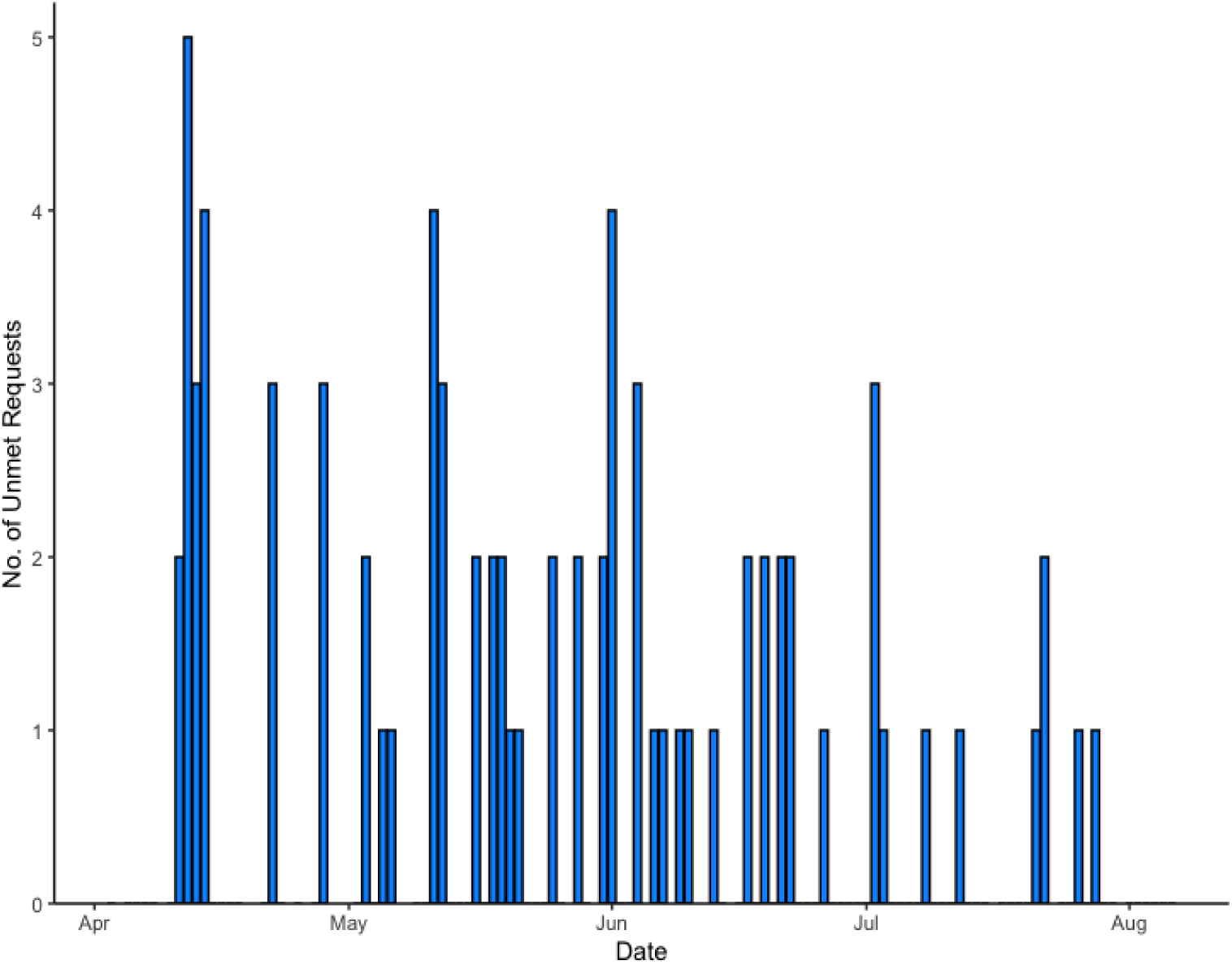
Number of unfilled requests per day. Each blue bar represents a day with a stockout, meaning there was no available tested blood in the blood bank at the hospital. Total number of days is 40 or 31.7% of the time.

A total of 531 whole blood units were collected from blood drives during this time; 149 (28.06%) units donated by females. Blood donations were mainly from community blood drives (333; 62.71%), but also consisted of replacement (150; 28.25%) and walk-in (48; 9.04%) donations (Supplemental Table 2).

### Laboratory Testing Data on RDT Safety

A total of 798 donors were tested with both RDT and EIA to assess the performance of RDT compared to EIA, the standard-of-care in Kenya. Overall TTI prevalence, via EIA quantification, was 5.4%. RDT diagnostic accuracy was compared to EIA (Table 3). The negative predictive value (NPV) for RDT was above 99% for each TTI analyte as well as collectively. There were no false positive results. There were seven negative RDT results that were positive by EIA. Repeat testing was performed on the seven false negatives (2 HIV, 1 HCV, 2 HBV, 2 Syphilis), and all cases were again negative by RDT.

**Table 2.** Themes from Qualitative Analysis.

|  | Theme | Quote |
| --- | --- | --- |
| 1 | Blood unavailability is untenable and must be overcome | “Sometimes we find we don’t have blood at all, so we are forced to just do what we can... start activating those people or those machineries that are responsible for blood in this hospital so that they help us out.”<br>“If there is a willing donor, that is something to consider; you do not [want to] let a patient die and there is someone here willing to donate for this patient”<br>“Those [hospital workers] are the immediate people who know how patients suffer, they feel that impact. You are handling a patient and the only thing they are lacking is they need blood and there is no otherwise, and you want to save the life of this patient, you just volunteer.” |
| 2 | Emergency measures using RDTs to expedite the issuance of blood products may be clinically necessary in limited circumstances | “I’ve actually had situations where I have had to do the rapid [test], crossmatch and give blood. I have had a couple of situations like that because there is no blood completely, even in maternity I have had to do that with [a] relative’s blood because I did not have blood. I have had to do that...several times.”<br>“So, most of the time when we are doing the rapid screening there is always that fear of maybe this is someone who is positive but there is seroconversion ... there is always that fear of maybe giving a blood that is not completely safe to the patient.”<br>“The policy is safe blood that we need to screen that blood and for it to be given a clean bill of health all quality control standards applicable ...but time after time we have had that situation where there is an emergency and we have blood, we decide to screen it ourselves and then as a control measure, the senior consultant there....must also sign against it.” |
| 3 | While a clinician must ultimately initiate and authorize an RDT-screened blood transfusion in times of dire emergency, the situation demands a shared burden of responsibility amongst the attending clinicians, patient and/or patient’s family | “It puts us in a situation of ethical dilemma because we know this is not supposed to be done, but then again, we know that we need to save life. It usually needs a team of people to think together on the way forward.”<br>“If the patient is awake, I can be able to converse with them, or if there is relative of sound mind you can share with them, give them the scenario, counsel them, give them the possible outcome if the patient does not get blood that is available...sometimes you can find it is possible for them to say no, now that one it will be difficult from my side considering there are medical-legal issues.”<br>“When the doctor has accepted to take responsibility.... we do the rapid test to save this life as long as the doctor has consented.” |
| 4 | The ideal donor is a voluntary, repeat donor, who is healthy, comes from a low-risk donor population, and has previously tested negative for transfusion transmitted infections, ideally through standard-of-care blood donor testing | “The regular blood donors come after three or four months.... normally we assume their blood is safe...at least that blood if you do a rapid and it turns out negative you can assume it is okay because this is someone who usually comes after every three or four months.”<br>“The first challenge of relatives and caretakers availability...they are available but they do not meet the criteria which is that they have to be old enough, they have to be physically fit, they have to be healthy.”<br>“The major challenge is that the blood needs to be taken ... for screening tests, and then you wait for the results so that they transfuse...it takes time.” |
| 5 | Supportive, comprehensive policy and consistent funding sources are necessary to improve blood availability and access in both standard and emergent settings | “[The] government should take initiative and make people donate daily [regularly], the process should be continuous, not seasonal because blood is ever needed in the hospital.”<br>“We need to develop...protocols to ensure the communication component...who needs what and at what time and what needs to be done, it is missing. It is just left to the proactiveness of the ward in charge or the clinician and the good relationship between the satellite blood bank team.”<br>“If we can have a testing lab within the hospital, I think that can really help us in terms of minimizing the time taken between donation and the time the blood is given to the patient.” |

RDT: Rapid diagnostic tests

**Table 3.** Diagnostic accuracy of RDTs compared against EIA standard. (n=798 samples)

|  | **HIV** | **CI** | **HBV** | **CI** | **HCV** | **CI** | **Syphilis** | **CI** | **All TTI** | **CI** |
| --- | --- | --- | --- | --- | --- | --- | --- | --- | --- | --- |
| Prevalence | 0.75%<br>(n=6) | 0-2% | 2.3% (n=18) | 1-4% | 0.75%<br>(n=6) | 0-2% | 2%<br>(n=16) | 1%-3% | 5.4%<br>(n=37) | 4%-7% |
| Sensitivity (SD) | 66% | 22%-96% | 94% | 73%-100% | 50% | 12%-88% | 81% | 54%-96% | 83.7% | 72%-95% |
| Specificity (SD) | 100% | - | 99.7% | 99%-100% | 100% | - | 100% | - | 100% | (0%) |
| Positive Predictive Value (SD) | 100% | (40%-100%) | 89.5% | (67%-99%) | 100% | (29%-100%) | 100% | (75%-100%) | 100% | (91%-100%) |
| Negative Predictive Value (SD) | 99.7% | (99%-100%) | 99.9% | (99%-100%) | 99.6% | (99%-100%) | 99.6% | (99%-100%) | 99.2% | (98%-100%) |

HBV = hepatitis B virus, HCV = hepatitis C virus, HIV = human immunodeficiency virus, SD = standard deviation, CI = confidence interval

Results from 1:1 RDT serial dilution along with associated viral loads and optical densities are displayed in Figure 2. HIV was consistently detectable to the 10th dilution; HBV to the 8^th^ dilution; HCV to the 3^rd^ dilution; and syphilis to the 7^th^ dilution.

**Figure 2.**
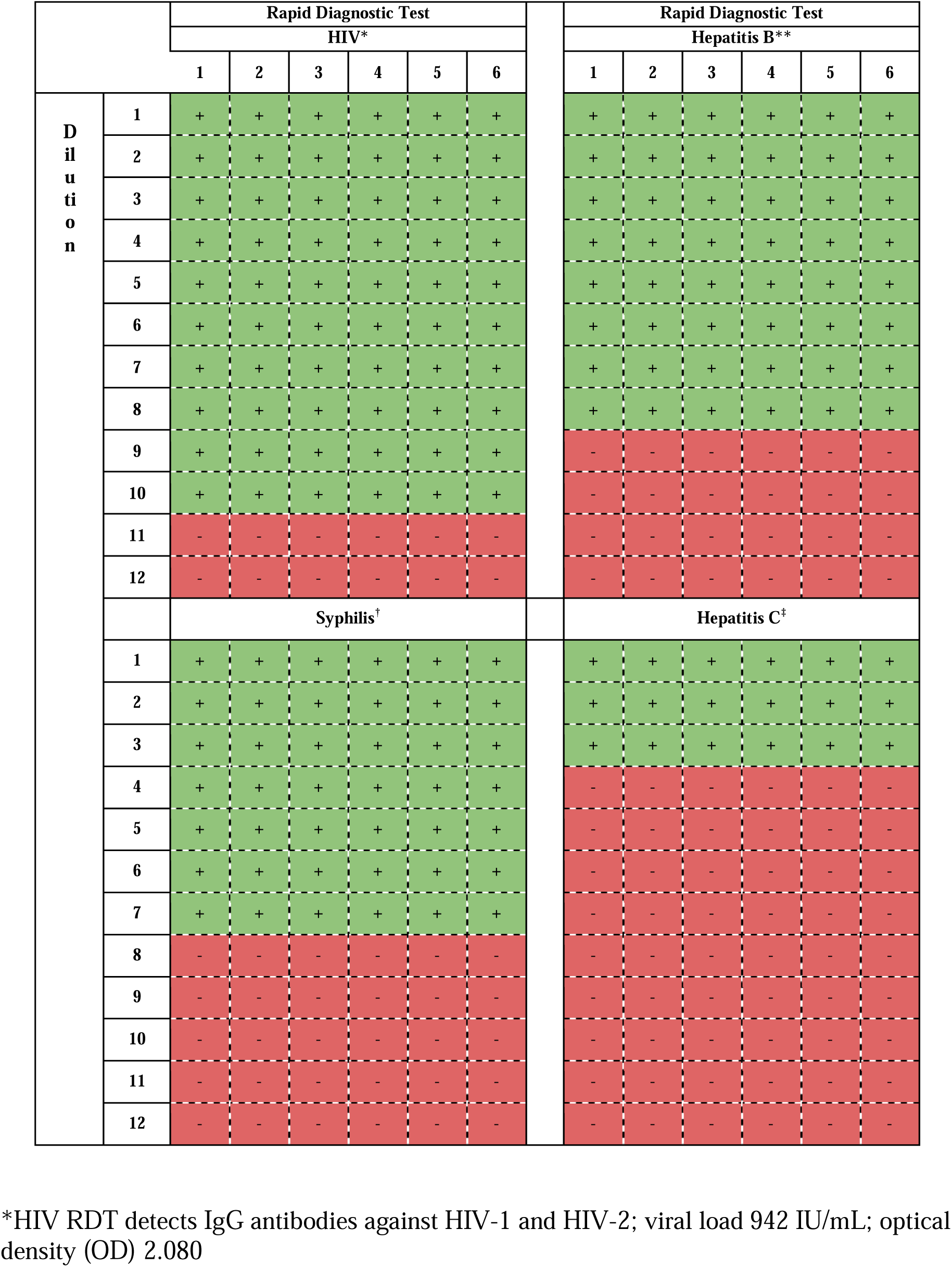
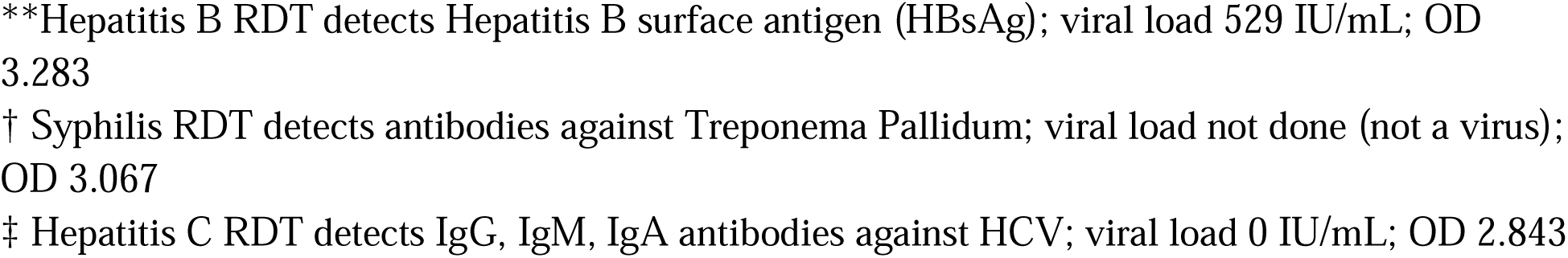
Serial dilution of RDT samples. Each sample was diluted in a 1:1 ratio with saline, where each subsequent dilution level is half the concentration of the previous level. RDT: rapid diagnostic tests.

### Qualitative Data on Need and Feasibility of Implementation

Interview participants included two county health administrators, the LCRH hospital administrator, two blood banking or hospital laboratory staff members, one nurse, and four clinicians (three medical officers, one clinical officer). Each key informant had between two to 20 years of practice experience.^25^ Five major themes emerged, encompassing perspectives around blood unavailability and just-in-time transfusion practices (see Table 2 for representative quotes, organized by theme):

#### 1. Blood unavailability is untenable and must be overcome

All participants reported situations where they had a patient whose survival depended on blood transfusion and compatible blood was unavailable. In some cases, compatible blood was present in the blood bank but awaiting TTI results from the RBTS, which could take 3-14 days. Staff members recounted contacting hospital facilities, hours away, for available units. Despite these efforts, situations arose where banked blood was unavailable in the timeframe needed and patients died or experienced severe complications as a result.

#### 2. Emergency measures using RDT to expedite issuance of blood products may be clinically necessary in specific circumstances

In extenuating circumstances – when a patient’s survival depended on blood transfusion, yet blood was unavailable – participants noted that using RDT screening to (1) facilitate transfusion from contemporaneous donor blood collection or (2) to screen stored blood pending EIA testing results in the blood bank, may be necessary. The only alternative for the patient, they noted, would be death. However, providers and local health system authorities were uncomfortable using RDT, expressing concerns about their legality of use for this purpose, the reliability of RDT performance, and the potential for TTI transmission. They also noted that using RDT in this circumstance – even if the purpose is to save a life – exposed providers to legal and moral risk if the survivor contracted a TTI.

#### 3. While clinicians must ultimately initiate and authorize RDT-screened blood transfusions, it demands shared responsibility amongst clinicians, hospital administration, and patients and/or their family

Participants emphasized the complexity of decision-making required to approve RDT-screened transfusions, considering medical risks to the patient as well as legal repercussions and moral injury to providers if TTI transmission occurred. Participants highlighted the importance of patient consent, i.e., patients should be well informed regarding the risks of RDT-screened transfusions. If patients are unable to consent, it must be obtained from family. These conversations should be led by clinicians after discussion with the blood bank confirming blood unavailability within the timeframe needed to save a patient’s life. Given the cooperation required among multiple stakeholders to facilitate this time-sensitive process, participants noted it was critical to have clear protocols and agreements in place beforehand to streamline communication and provide institutional support for clinical decisions made by staff. Participants specified that the burden of making these decisions should not be borne solely by the clinical providers who authorize the decisions but should be shared by the entirety of the hospital system.

#### 4. The ideal donor is a voluntary, healthy, repeat donor, who comes from a low-risk donor population, and has previously tested negative for TTIs, ideally through standard-of-care testing

When the clinical team has decided to proceed with emergency RDT-based transfusion, participants noted it is essential that all efforts be made to minimize risks of transfusion-related illness. The ideal donor, they note, has previously donated, with updated TTI screening and health information on file. These factors instill higher confidence in RDT results. However, finding such donors in a short time frame may be prohibitively challenging, and it is essential to use a stepwise approach to find donors. Participants indicated healthcare workers at LCRH are frequently called upon for this purpose and are considered relatively low risk because of their medical understanding and perceived likelihood of avoiding high-risk behaviors. Moreover, they are usually available and willing to donate in times of emergency. Nonetheless, participants also indicated that in the most emergent situations, the most available potential donor was accompanying family. They repeatedly emphasized the higher risk of relying on family donors and the preference to exhaust other opportunities before proceeding to this step. Relatives are often first-time donors, with varying degrees of knowledge regarding their health and highly motivated to donate for loved ones. In these situations, clinical risk of delayed transfusion must be weighed against the risk of potentially harmful transfusions.

#### 5. Supportive, comprehensive policy and consistent funding sources are necessary to improve blood availability and access

While participants agreed that circumstances do occur where emergency RDT-based transfusion processes are necessary, all emphasized the need for long-term, sustainable approaches to strengthen existing blood banking systems. Many reported the need for more support in conducting community blood drives, including reliable transportation and year-round funding. They also expressed frustration regarding current processes for testing blood at centralized laboratories, which create severe delays in transfusions. Many participants felt having equipment on site would streamline the process, given LCRH already collects and stores blood.

## Discussion

This is one of the few studies that specifically characterizes blood availability and insufficiency at a facility in a blood desert, and it reveals a profound shortage of blood at LCRH.^26^ Over a four-month study period, there were only 485 documented transfusion requests at LCRH, the core referral hospital for a population of 1.2 million people. This represents less than 10% of actual blood need annually based on models of population size and disease burden, recognizing that not all patients in need of blood can reach LCRH, a percentage of the population will seek care at other facilities, and providers may order less blood knowing that blood is not available. Nonetheless, despite this depressed demand, only 405 units of blood (85%) were transfused over the same study period. Stockouts – when unmet requests exceeded met requests on a given day – occurred during almost a third (31%) of the study period. During each stockout day, the number of unmet requests ranged from 1 to 5 units (Figure 1). Put in context, if a patient were to present at LCRH with life-threatening hemorrhage or severe anemia on these days, they would be at risk of death from lack of transfusion.

Faced with such circumstances, providers reported already using an RDT-based emergency transfusion process – effectively a walking blood bank – in cases of extreme emergency and advocated its continued use in emergencies when banked, screened blood was unavailable. Providers felt obligated to use emergency measures to save life and reported using RDT to screen freshly collected blood or previously collected blood awaiting screening results from the RBTS for the purposes of transfusion. Meanwhile, participants remained concerned that RDT had substandard performance compared to standard-of-care EIA testing, and that they were exposing patients to potential TTI transmission and themselves to potential legal liability. However, participants underscored the moral burden of knowing donors and blood were available while patients were on the verge of death from hemorrhage or anemia, and the potential for TTI transmission, versus the finality of death from lack of transfusion. “*We cannot let the patient die when a willing donor is present.*”

Fortunately, in our safety testing comparing RDT performance against EIA in real-world conditions of Turkana County, RDT performed extremely well even in the scorching desert heat and extended transport times for consumables. If blood samples collected in Lodwar screened negative for TTIs using RDTs, there was a 99.2% likelihood they would also screen negative with standard-of-care testing. There were no false positives; RDT showed excellent consistency with no false negatives in serial dilution assays until below the limit of detection. To reiterate, all RDT in this study was procured, stored, and used under standard institutional processes and conditions, with testing performed by LCRH personnel.

Together, we believe these findings add to the compelling arguments for WBBs as a stopgap measure to address the crisis of death from hemorrhage and severe anemia when banked blood is unavailable, and especially for blood desert contexts like Turkana County where banked blood is rarely available.^5^ Despite decades of concern about RDT reliability for transfusion safety, our data demonstrates excellent negative predictive value in this setting.^27^

Of note, our findings have important limitations. This was a single center study that housed a functional blood bank with the only major deficit being the inability to perform standard-of-care EIA testing on-site. In other words, the WBB model was deemed feasible by providers and a hospital system that was capable of mobilizing blood donors, collecting blood, performing critical crossmatch testing, and administering transfusion. Not all facilities in blood desert contexts have this institutional skillset and further research is urgently needed to evaluate translation of these findings to these settings. Additionally, the NPV of 99.2% for RDT compared to EIA is dependent on the local prevalence rates for TTIs in the population. In our study, the baseline rate of TTI in our donor population was 5.4%, likely influenced by strong donor pre-screening protocols. Where endemic TTI rates are lower, the NPV will be even higher; where TTI rates are higher, the NPV will decrease. Stakeholders must take local rates of TTI into context when implementing RDT-based emergency transfusion protocols. Finally, our study was underpowered to assess additional test characteristics (e.g. sensitivity) of RDT performance for specific TTIs given the relatively small difference in test performance between RDT and EIA. Nevertheless, it was powered to determine NPV performance, the most critical test characteristic when considering whether to proceed with emergency transfusion, and we were able to test almost 800 samples over six months with wide donor variation.

Validated models in a range of clinical environments are needed to establish RDT performance in additional settings. High quality implementation studies are needed to establish standardized, yet context-specific approaches for WBB activation, donor mobilization, testing policies, training of professionals, and high-quality approval and review processes to minimize risks to the patient and provider. A WBB can be a stopgap measure to address the extensive burden of hemorrhagic shock and severe anemia in the world’s lowest resource civilian settings, where patients and their providers struggle without timely access to blood through a blood bank.

## Supporting information

LBP1 Supp Table 1

LBP1 Supp Table 2

LBP1 Supp 3

## Data Availability

All data produced in the present study are available upon reasonable request to the authors

## Acknowledgements

Funding for this project was provided by grant support from the Gillian Reny Stepping Strong Center for Trauma Innovation. Project execution was made possible through resource sharing and collaboration with the Pathways for Innovative Transfusion Systems-Kenya Study, National Institutes of Health National Heart, Lung, Blood Institute BLOODSAFE Initiative. We thank the Turkana County government, the leadership and staff of the Lodwar County Referral Hospital, and the Kenya Tissue and Transplant Authority for their critical collaboration.

## Author Contributions

NPR, TC, and LSB conceptualized the study, applied for funding, and supervised the project. NK coordinated study materials and led the writing along with NPR, TK, and LSB. NR, TC, LSB, NK, CM, AM, WW, SO, EM, VR, AMV conducted the investigation and data curation. NPR, TC, LSB, WW, APC, MDS, TD, SG, PK, SK, JCP, VR, NR, NT, AT, RS, ES, FM, and AMV contributed to qualitative methodology, analysis, and writing. NK and TD performed formal quantitative analysis. MD advised on quantitative analysis for RDT testing. JCP, PK, CW, NR, WW consulted on study design and execution. All authors reviewed and approved the final manuscript.

