## Supplementary material for "The Local Initiative For Emergency Blood (LIFE-Blood) Study: A mixed-methods, single center exploration of civilian walking blood bank need, feasibility, and safety in a low-resource blood desert": LBP1 Supp Table 1

**Supplemental Table 1.** The number of requests for whole blood by hospital ward and the number filled by the blood bank.

| Ward | Total Requests | Requests Filled | |
|---|---|---|---|
| | | Yes | No |
| **Surgical** | 146 (30.04%) | 122 (83.56%) | 24 (16.44%) |
| **Medical** | 134 (27.57%) | 118 (88.06%) | 16 (11.94%) |
| **Maternity** | 87 (17.90%) | 71 (81.61%) | 16 (18.39%) |
| **Pediatrics** | 72 (14.81%) | 60 (83.33%) | 12 (16.67%) |
| **Accident & Emergency** | 21 (4.32%) | 18 (85.71%) | 3 (14.29%) |
| **ICU** | 19 (3.91%) | 14 (73.68%) | 5 (26.32%) |
| **Operating Theatre** | 3 (0.62%) | 2 (66.67%) | 1 (33.33%) |
| **Gynecology** | 2 (0.42%) | 2 (100%) | 0 |
| **HDU** | 1 (0.21%) | 1 (100%) | 0 |
| **Oncology** | 1 (0.21%) | 1 (100%) | 0 |
| **Total** | 486 (100%) | 409 (84.16%) | 77 (15.84%) |

HDU = High Dependency Unit, ICU = Intensive Care Unit
