## Supplementary material for "The Local Initiative For Emergency Blood (LIFE-Blood) Study: A mixed-methods, single center exploration of civilian walking blood bank need, feasibility, and safety in a low-resource blood desert": LBP1 Supp Table 2

**Supplemental Table 2. Demographics of blood donors at LCRH from April to July 2022.**

| Month | Total Donors | Female (%) | Total Replacement Donors | Female (%) | Total Walk-in Donors | Female (%) | Total Drive Donors | Female (%) |
|---|---|---|---|---|---|---|---|---|
| Apr | 69 | 19 (27) | 54 | 14 (25) | 15 | 5 (33) | 0 | 0 (0) |
| May | 121 | 21 (17) | 29 | 9 (31) | 10 | 3 (30) | 82 | 9 (10) |
| June | 224 | 79 (35) | 30 | 0 (0) | 15 | 4 (26) | 179 | 75 (41) |
| July | 117 | 30 (25) | 37 | 6 (16) | 8 | 1 (12) | 72 | 23 (31) |
| TOTAL | 531 | 149 (28) | 150 | 29 (19) | 48 | 13 (27) | 333 | 107 (32) |
