## Supplementary material for "The Local Initiative For Emergency Blood (LIFE-Blood) Study: A mixed-methods, single center exploration of civilian walking blood bank need, feasibility, and safety in a low-resource blood desert": LBP1 Supp 3

**Supplemental 3.** Real-world patient scenario prompts

Scenario One

A man between the ages of 25-30 years old presents after a motorcycle collision in Lodwar. He is actively bleeding and the clinician orders blood. The clinician anticipates demise within 4 hours without blood. There is matching blood in the Lodwar satellite blood center, but it has not received screening test results from Regional Blood Transfusion Center-Eldoret.

Scenario Two

A woman between the ages of 20-25 years old is postpartum day 2 referred from another village with severe anemia and active bleeding from postpartum hemorrhage. The clinician anticipates demise within 4 hours without blood. There are no matching blood units in the Lodwar satellite blood center. She is accompanied by a family member.
